# Correlation of SARS-CoV-2 nucleocapsid antigen and RNA concentrations in nasopharyngeal samples from children and adults using an ultrasensitive and quantitative antigen assay

**DOI:** 10.1101/2020.11.10.20227371

**Authors:** Nira R. Pollock, Timothy J. Savage, Hanna Wardell, Rose Lee, Anu Mathew, Martin Stengelin, George B. Sigal

## Abstract

**Background:** Diagnosis of COVID-19 by PCR offers high sensitivity, but the utility of detecting samples with high cycle threshold (Ct) values remains controversial. Currently available rapid diagnostic tests (RDTs) for SARS-CoV-2 nucleocapsid antigens (Ag) have sensitivity well below PCR. The correlation of Ag and RNA quantities in clinical nasopharyngeal (NP) samples is unknown.

**Methods:** An ultrasensitive, quantitative electrochemiluminescence immunoassay for SARS-CoV-2 nucleocapsid (the MSD^®^ S-PLEX^®^ CoV-2 N assay) was used to measure Ag in clinical NP samples from adults and children previously tested by PCR.

**Results:** The S-PLEX Ag assay had a limit of detection (LOD) of 0.16 pg/mL and a cutoff of 0.32 pg/mL. Ag concentrations measured in clinical NP samples (collected in 3.0 mL media) ranged from less than 160 fg/mL to 2.7 ug/mL. Log-transformed Ag concentrations correlated tightly with Ct values. In 35 adult and 101 pediatric PCR-positive samples, sensitivity was 91% (95% CI, 77-98%) and 79% (70-87%), respectively. In samples with Ct ≤ 35, sensitivity was 100% (88-100%) and 96% (88-99%), respectively. In 50 adult and 40 pediatric PCR-negative specimens, specificity was 100% (93-100%) and 98% (87-100%), respectively.

**Conclusions:** Nucleocapsid concentrations in clinical NP samples span 8 orders of magnitude and correlate closely with RNA concentrations (Ct values). The S-PLEX Ag assay had 96-100% sensitivity in samples from children and adults with Ct values ≤ 35, and 98-100% specificity. These results clarify Ag concentration distributions in clinical samples, providing insight into the performance of Ag RDTs and offering a new approach to diagnosis of COVID-19.

**Key points:** SARS-CoV-2 nucleocapsid concentrations in clinical nasopharyngeal samples, measured with an ultrasensitive assay, spanned an 8-log range and correlated closely with PCR Ct values. The assay was 96-100% sensitive in pediatric/adult samples with Ct values ≤ 35, and 98-100% specific.

## Introduction

Nucleic acid amplification tests (NAAT) for SARS-CoV-2, the etiologic agent of COVID-19, can be highly sensitive for diagnosis of COVID-19 and are being performed in centralized laboratories around the globe in staggering numbers [1, 2]. However, NAAT capacity and utility have been impacted world-wide by recurring shortages of testing reagents and logistic barriers causing long delays in results turn-around time. The need for more rapid and decentralized testing options has led to swift development of rapid diagnostic tests (RDTs) for point-of-care (POC) use that detect SARS-CoV-2 nucleocapsid antigen (Ag) in as little as 15 minutes. Reported clinical sensitivities for Ag RDTs vary widely, ranging from 74 to 97% versus PCR [3-7] when performed at POC in patients suspected of COVID. Concerns about both false negative and false positive Ag RDT results [8] have raised caution regarding implementation in many settings in which their use is being considered, including K-12 schools, nursing homes, and community testing centers.

In symptomatic adults, viral loads in nasopharyngeal (NP) samples (as measured by NAAT) rise approximately two days prior to symptom onset, peak in 2-4 days, remain high over the first 7 days of symptoms, then decrease to undetectable levels over a variable time frame. RNA levels in asymptomatically infected adults appear to follow similar kinetics [9, 10]. However, the time course for Ag concentration is not as well understood due to the lack of highly sensitive and quantitative assays for Ag measurement.

We have developed quantitative immunoassays for SARS-CoV-2 nucleocapsid Ag using conventional R-PLEX^®^ and enhanced ultrasensitive S-PLEX electrochemiluminescence (ECL) assay formats [Meso Scale Discovery^®^ (MSD)] and used these assays to measure Ag concentration in clinical NP swab eluates previously tested by PCR for clinical diagnosis. Our goal was to assess Ag concentration distributions in clinical samples, correlate Ag concentrations with Ct values as a measure of RNA concentration, and assess the sensitivity and specificity of the S-PLEX Ag assay for detection of SARS-CoV-2 in clinical samples versus PCR as a reference method.

## Methods

### Immunoassay Protocols

ECL immunoassays for SARS-CoV-2 nucleocapsid Ag, employing sandwich immunoassay formats, were carried out using assay kits, instrumentation and multi-well plate consumables from MSD [11]. The multi-well plate consumables have integrated screen-printed carbon ink electrodes on the bottom of each well that are used as solid-phase supports for binding reactions, and as the source of electrical energy for inducing ECL from ECL labels in binding complexes on their surfaces. Two ECL assay formats were employed: a conventional ECL format (the MSD R-PLEX SARS CoV-2 N assay kit run on MSD U-PLEX^®^ plates [12] and an ultrasensitive ECL format [13-15] that provides additional signal enhancement and sensitivity (the MSD S-PLEX CoV-2 N assay kit). The assays were run according to protocols that are provided in the kit package inserts. Details of the immunoassay protocol, quantitation, and statistical analysis are provided in Supplementary Methods.

### Testing of Nasopharyngeal Swab Samples

NP swab samples in universal transport media or saline were tested in duplicate. Concentrations were calculated for each replicate and then averaged. Samples with calculated concentrations below the LOD or above the top calibrator were assigned the LOD or top calibrator values, respectively, for statistical analyses and graphing. Samples were retested if they provided a signal above the assay threshold and the coefficient of variation (CV) for the replicate measurements was greater than 25%, or if there was a reported operator error during processing. Samples that provided a calculated concentration greater than 50% of the top calibrator in the R-PLEX assay were rerun in the R-PLEX assay at a 1:100 dilution. To characterize the distribution of nucleocapsid concentrations and their correlation to PCR Ct values, we used the following approach to assign a concentration value to each sample, given that the range of concentrations was greater than the dynamic range of either assay: (i) we used the concentration measured using the S-PLEX assay if it was less than half the concentration of the top S-PLEX calibrator, (ii) otherwise, we used the concentration measured using the R-PLEX assay if it was less than half the concentration of the top R-PLEX calibrator, (iii) otherwise, we used the concentration measured using the R-PLEX assay for a 1:100 diluted sample.

### Analysis of a Viral Culture Preparation

The concentration of nucleocapsid was measured in a reference cell culture preparation of inactivated (gamma irradiation) SAR-CoV-2 virus (Isolate USA-WA1/2020, Catalog Number NR-52287, BEI Resources) with assigned values for the concentration of infectious virus (2.8 x 10^5^ TCID50/mL prior to inactivation) and RNA (4.1 x 10^9^ copies/mL). Serial dilutions that provided concentrations in the quantitation range of the assay were corrected for dilution then averaged to determine the concentration of nucleocapsid in the sample. Assay cross-reactivity for circulating coronaviruses OC43 and 229E was measured in untitered viral culture supernatants (diluted 1:100) obtained from ATCC (catalog numbers VR-740 and VR-1558, respectively).

### Clinical samples

All samples were clinical NP swab samples eluted in either 3.0 mL Copan Universal Transport Medium (Copan, Murietta, CA) or 3.0 mL normal saline (Remel, Lenexa, KS). Samples were obtained from adult hospital employees or pediatric patients undergoing clinical testing for suspected SARS-CoV-2. Clinical data was obtained by chart review.

The majority of the samples were tested by the Hologic Panther Fusion assay (Hologic, San Diego, CA); 2 adult samples were tested by a laboratory-developed test utilizing altona Diagnostics reagents [16]. The majority of PCR results were reported approximately 4 hours after sample collection and residual volumes were aliquoted and frozen (−80C) shortly afterwards. Deidentified aliquots were shipped frozen to MSD for testing.

This study was performed under approval from the Boston Children’s Hospital IRB with waiver of informed consent; only fully deidentified data were analyzed.

## Results

### Assay analytical performance

The calculated LODs for the R-PLEX and S-PLEX Ag assays were 2 pg/mL and 0.16 pg/mL, respectively. Based on these values, we set the assay thresholds for classifying positive samples as 2 x LOD or 4 pg/mL and 0.32 pg/mL, respectively. Nucleocapsid concentration in an irradiated SARS-CoV-2 culture preparation (BEI, Methods), as measured using the S-PLEX Ag assay, was compared to the concentrations provided by BEI in TCID50 and RNA copies per mL. We calculated the ratio of Ag to virion as 0.89 pg per TCID50 (or 1.1 x 10^7^ protein molecules per TCID50) and to RNA as 1.5 x 10^−4^ pg per RNA copy. This ratio corresponds to about 1.8 x 10^3^ protein molecules per RNA copy, based on a molecular weight of 49 kD for SARS-CoV-2 nucleocapsid, which is higher than the 200 to 400 nucleocapsid molecules per genome in a typical coronavirus particle [17, 18]. Using these conversion factors, virus concentrations measured at the Ag assay thresholds are around 4.5 TCID50/mL and 27,000 RNA copies/mL for the R-PLEX assay and 0.36 TCID50/mL and 2,100 RNA copies/mL for the S-PLEX assay. The analytical sensitivity of the S-PLEX Ag assay is, therefore, on par with the LODs of many PCR assays (2 copies/uL sample)[19]. In comparison, the detection limits for the commercial BD Veritor and Quidel Sofia SARS-CoV-2 antigen tests, based on the values in the package inserts, are roughly 140 TCID50/mL and 226 TCID50/mL, which based on our calculated conversion factors correspond to 121 pg/mL and 204 pg/mL of Ag, respectively, or roughly 500-fold less sensitive than the S-PLEX Ag test.

Assay specificity was confirmed by testing culture preparations of the 229E and OC43 circulating coronavirus strains. No measurable cross-reactivity was observed with these strains. The assay was highly reproducible. CVs for the low and high concentration controls run in duplicate on each assay plate during the study (10 plates per assay, run over several days) were 7.7% and 8.3%, respectively, for the R-PLEX assay and 7.0% and 7.7% for the S-PLEX assay.

### Distribution of nucleocapsid antigen concentrations in clinical samples

Nucleocapsid concentrations were measured in NP samples from 85 adult employees (35 PCR positive, 50 PCR negative) and 141 pediatric patients (101 PCR positive, 40 PCR negative). Each sample was the earliest available sampling time-point for that employee/patient in the hospital system. The clinical characteristics of the adult and pediatric patients are summarized in Table 1. 4/35 PCR-positive adult employees and 2/101 PCR-positive pediatric patients were asymptomatic at the time of testing.

The measured concentrations of nucleocapsid in the swab samples, as measured using the R-PLEX and S-PLEX assays without any additional sample dilution, are shown in Supplementary Figure 1. Using the optimal assay format or dilution for each sample (Methods), Figure 1 plots the distributions of Ag concentrations in adult and pediatric samples that tested negative or positive by PCR. Figure 1 also shows the assay thresholds for the S-PLEX and R-PLEX nucleocapsid assays, and for comparison, the estimated LODs for the Quidel Sofia and BD Veritor assays. PCR-negative samples clustered tightly with measured Ag concentrations around or below the LOD for the S-PLEX assay, and only one sample providing a concentration slightly above the S-PLEX assay threshold. Ag concentrations for PCR-positive samples were evenly distributed over a wide range of concentrations spanning almost 8 orders of magnitude (from < 160 fg/mL to 2.7 ug/mL) in samples from both adults and children. The sample incubation step in the assay protocol includes 0.5% Triton X-100 to release Ag from virus particles. However, testing of a small set of samples without Triton generated similar calculated concentrations, suggesting that most of the Ag in the sample was not confined within intact virus particles.

**Figure 1.**
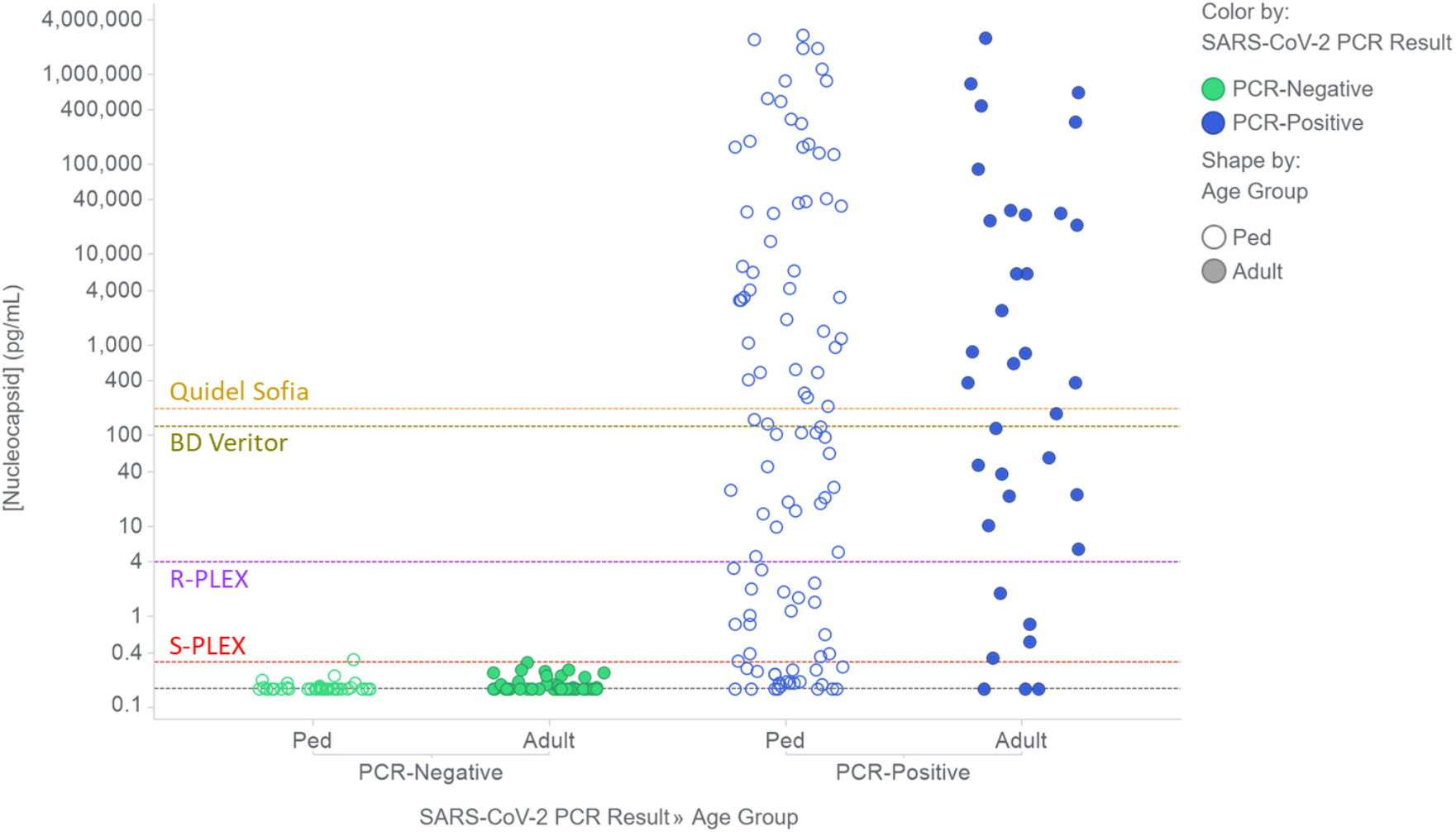
Nucleocapsid concentrations in clinical NP swab samples from PCR-negative (green) and PCR-positive (blue) adults (filled circles) and children (Ped, open circles). The figure shows the concentrations measured using the optimal assay format and dilution for each sample as described in the Methods section. Concentrations below the LOD for the S-PLEX assay were assigned the LOD value (grey dashed line). The plots also show dashed lines to indicate the applied assay thresholds for the ultrasensitive S-PLEX and conventional R-PLEX ECL assays, as well as the estimated analogous values for the commercial BD Veritor and Quidel Sofia systems.

Figure 2 plots the measured Ag concentrations for the PCR-positive samples from Figure 1 against the PCR Ct values for those samples. Linear regression demonstrates a strong (R^2^ = 0.90) correlation of the log2-transformed concentration values (in pg/mL) with Ct values across the large range of values measured with both assays. The estimates (and 95%CIs) for the slope and y-intercept are −0.87 (−0.92 to −0.82) and 30.9 (29.5 to 32.2). Given that a one unit decrease in Ct value should correspond to just under a doubling of the RNA concentration, a slope for the relationship of log2 nucleocapsid concentration with Ct that is close to −1.0 is consistent with a roughly linear relationship between viral protein and RNA concentrations. Using the regression model, the expected Ct values for samples at the antigen assay thresholds for the R-PLEX and S-PLEX ECL Ag assays, and the commercial BD and Quidel assays are 34, 38, 28 and 27 cycles respectively.

**Figure 2.**
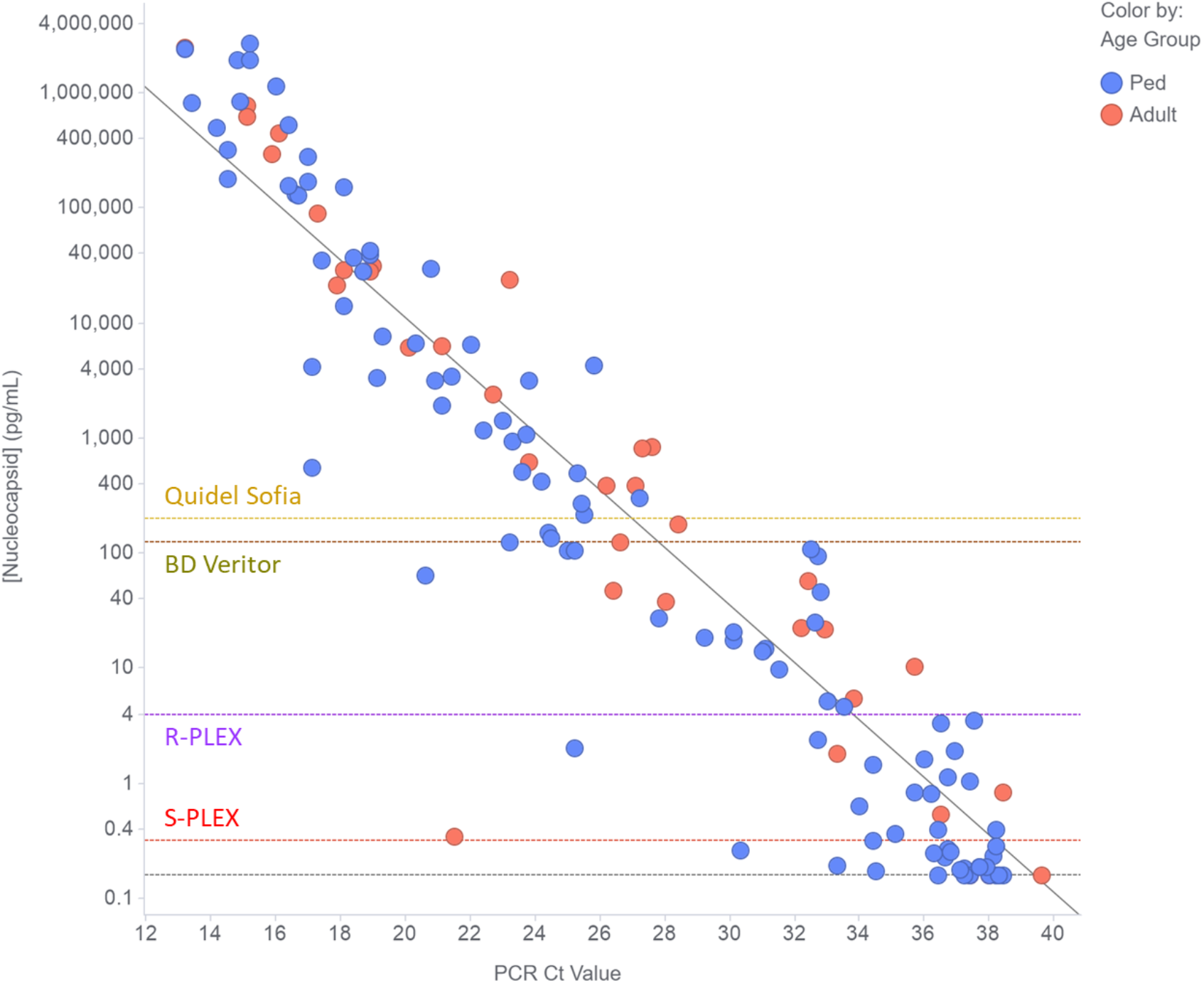
Correlation of nucleocapsid concentration with PCR Ct value for clinical NP swab samples from PCR-positive adults (red circles) and children (Ped, blue circles). The figure shows the concentrations measured using the optimal assay format and dilution for each sample as described in the Methods section. Concentrations below the LOD for the S-PLEX assay were assigned the LOD value (grey dashed horizontal line). The diagonal dashed grey line is the linear regression fit to the data (using log-transformed nucleocapsid concentrations). To provide estimates of the expected Ct value for samples at the threshold for different nucleocapsid assay formats, the plots also show horizontal dashed lines to indicate the applied assay thresholds for the ultrasensitive S-PLEX and conventional R-PLEX ECL assays, as well as the estimated analogous values for the commercial BD Veritor and Quidel Sofia systems.

### Assay clinical performance

The sensitivity and specificity of the S-PLEX Ag assay versus PCR in adult and pediatric samples are shown in Table 2. Specificity in PCR-negative samples was 100% (93-100%) and 98% (87-100%) in adult and pediatric samples, respectively. Sensitivity in PCR-positive samples was 91% (77%-98%) and 79% (70%-87%) in adult and pediatric samples, respectively. The correlation plot in Figure 2 shows that the lower sensitivity in pediatric samples is largely accounted for by a number of pediatric samples with Ct values between about 36 and 38 (Supplementary Table 1). Considering only samples with Ct values ≤ 35, the S-PLEX Ag test sensitivity increased to 100% (88-100%) and 96% (88-99%), respectively, for adult and pediatric samples. We note that these high-Ct samples are also potentially on the borderline for detection by the Hologic assay, as the Ct value corresponding to the Panther Fusion LOD is 35.6 per the package insert [20].

**Table 1.**
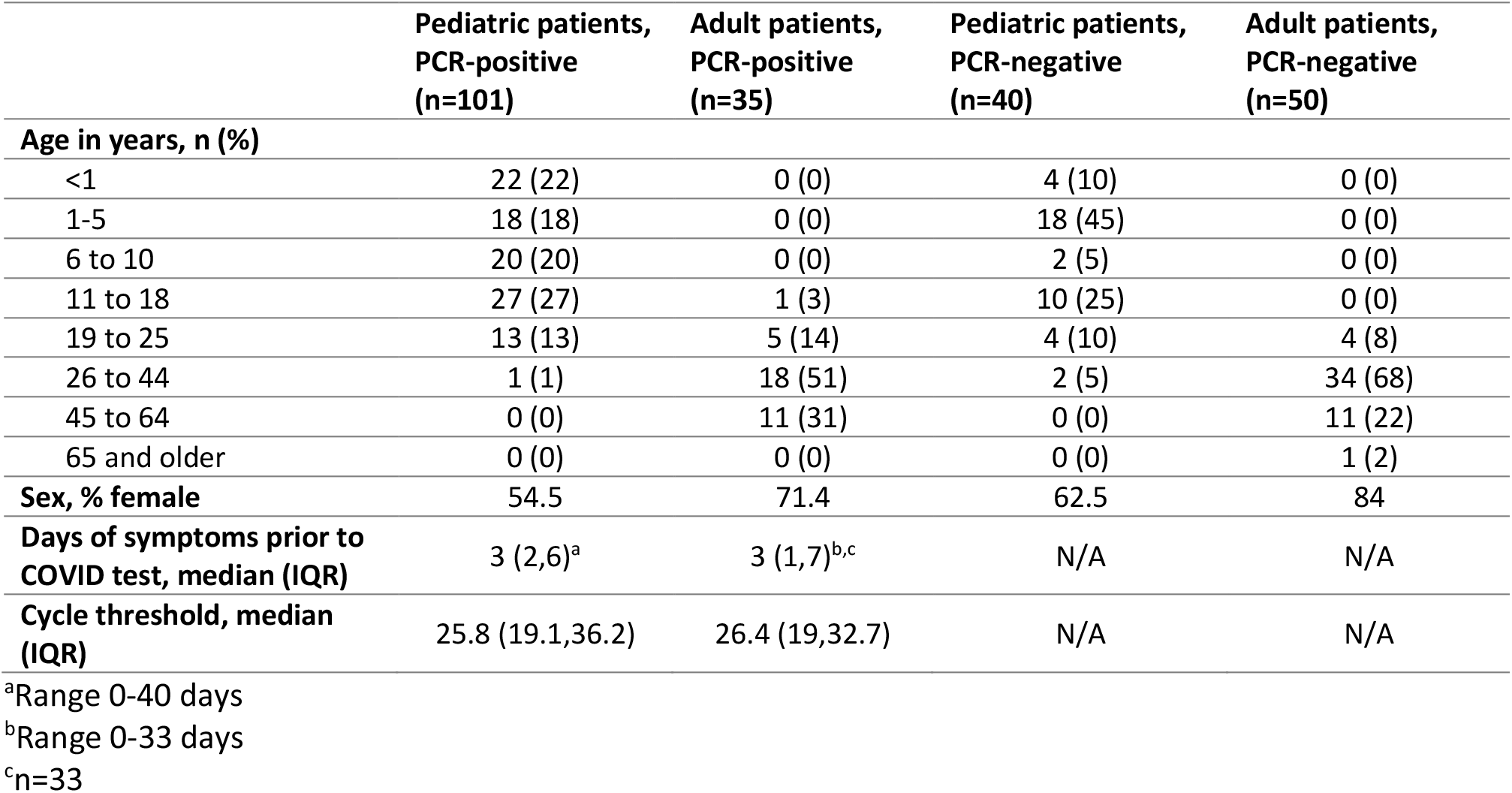
Clinical and laboratory characteristics of adult and pediatric patients contributing samples.

|  | Pediatric patients,<br>PCR-positive<br>(n=101) | Adult patients,<br>PCR-positive<br>(n=35) | Pediatric patients,<br>PCR-negative<br>(n=40) | Adult patients,<br>PCR-negative<br>(n=50) |
| --- | --- | --- | --- | --- |
| **Age in years, n (%)** |  |  |  |  |
| <1 | 22 (22) | 0 (0) | 4 (10) | 0 (0) |
| 1-5 | 18 (18) | 0 (0) | 18 (45) | 0 (0) |
| 6 to 10 | 20 (20) | 0 (0) | 2 (5) | 0 (0) |
| 11 to 18 | 27 (27) | 1 (3) | 10 (25) | 0 (0) |
| 19 to 25 | 13 (13) | 5 (14) | 4 (10) | 4 (8) |
| 26 to 44 | 1 (1) | 18 (51) | 2 (5) | 34 (68) |
| 45 to 64 | 0 (0) | 11 (31) | 0 (0) | 11 (22) |
| 65 and older | 0 (0) | 0 (0) | 0 (0) | 1 (2) |
| **Sex, % female** | 54.5 | 71.4 | 62.5 | 84 |
| **Days of symptoms prior to**<br>**COVID test, median (IQR)** | 3 (2,6)<sup>a</sup> | 3 (1,7)<sup>b,c</sup> | N/A | N/A |
| **Cycle threshold, median**<br>**(IQR)** | 25.8 (19.1,36.2) | 26.4 (19,32.7) | N/A | N/A |

<sup>a</sup>Range 0-40 days
<sup>b</sup>Range 0-33 days
<sup>c</sup>n=33

**Table 2.** **Performance of the S-PLEX Ag assay versus PCR for detection of SARS-CoV-2 in samples from adult and pediatric patients**

|  | PCR-Negative Patients |  | PCR-Positive Patients<br>All Ct Values |  | Ct Values ≤ 35 |  |
| --- | --- | --- | --- | --- | --- | --- |
|  | N | Specificity (95% CI) | N | Sensitivity (95%CI) | N | Sensitivity (95%CI) |
| **Age Group** |  |  |  |  |  |  |
| Pediatric | 40 | 98% (87%-100%) | 101 | 79% (70%-87%) | 72 | 96% (88%-99%) |
| Adult | 50 | 100% (93%-100%) | 35 | 91% (77%-98%) | 29 | 100% (88%-100%) |
| Combined | 90 | 99% (94%-100%) | 136 | 82% (75%-88%) | 101 | 97% (92%-99%) |

This data set of measured nucleocapsid values provides a tool for estimating the clinical sensitivity that can be achieved with less sensitive assays. Based on the S-PLEX assay measurements, the sensitivity of the R-PLEX ECL assay can be predicted based on its 4 pg/mL threshold to be 68%, which agrees well with the actual observed sensitivity for this assay of 70% (Supplementary Table 2) based mostly on measurements with the S-PLEX assay. Using the estimated LODs for the BD and Quidel assays, the predicted sensitivities of the commercial assays for this sample set would be approximately 51% and 47%, respectively; sensitivity in samples with Ct < 30 would be 88% and 82% and with Ct < 25 would be 97% and 92%, respectively.

### Characterization of discordant samples

Samples with discordant PCR and S-PLEX Ag assay results were analyzed further. 21/24 false negative samples were from pediatric patients, and 21/24 had Ct values >35 (Supplementary Table 1). Since the linear regression model relating nucleocapsid concentration with Ct value predicted measurable nucleocapsid levels up to Ct values of 38, we conducted spike recovery experiments (Supplementary Methods) to determine if matrix interference may have led to under-detection of antigen. Only 3/24 of the samples exhibited significant matrix interference (spike recovery < 25%).

### Relationship between nucleocapsid concentration and symptom duration

The relationship between nucleocapsid concentration and PCR Ct value as a function of duration of illness (defined as days after onset of symptoms) was evaluated by linear regression (Figure 3). Due to the wide spread in Ag concentrations and Ct values, the correlations were relatively weak [R2 values were 0.079 and 0.074 for nucleocapsid (after log2-transformation) and PCR Ct value, respectively, although as expected both nucleocapsid and RNA concentrations decreased on average with time. Notably, the slope of decline was similar for the two analytes (Fig 3).

**Figure 3.**
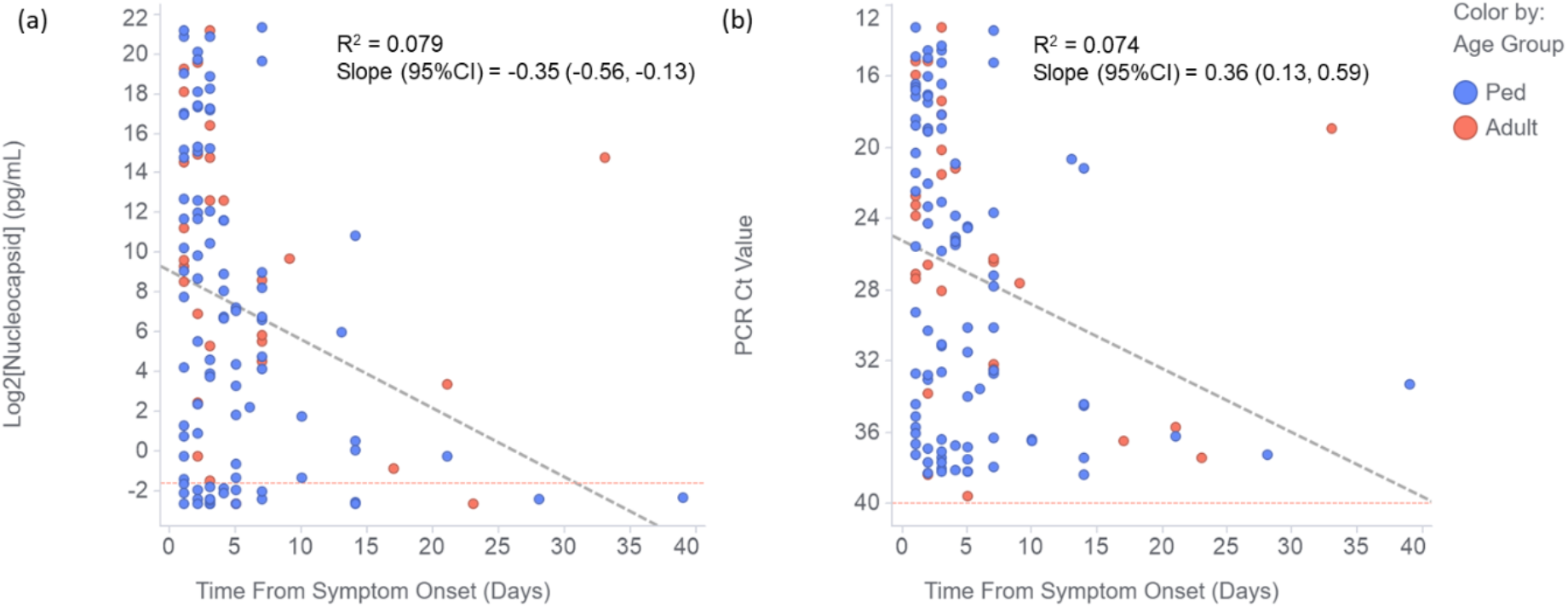
Correlation of (a) nucleocapsid concentration (log2-tranformed) and (b) PCR Ct value with time from symptom onset for clinical NP swab samples from PCR-positive adults (red circles) and children (Ped, blue circles). The plots also show the linear regression fit to the data (diagonal dashed grey lines).

## Discussion

This is the first characterization of SARS-CoV-2 nucleocapsid antigen concentration distributions in clinical nasopharyngeal swab samples. We compared measured Ag concentrations and qualitative (positive/negative) results to PCR results and Ct values generated using the Hologic Panther Fusion test, which has one of the lowest LODs among molecular assays tested with an FDA reference panel [19]. Our work has demonstrated that Ag concentrations in clinical samples span 8 orders of magnitude and that there is a close correlation between Ag and RNA concentrations (as reflected by Ct values) throughout the entire range of viral loads in adult and pediatric individuals with a range of symptom duration prior to testing. The S-PLEX Ag assay had near perfect specificity and high sensitivity compared to PCR, with near 100% sensitivity in NP swab samples with Ct values ≤ 35, offering the potential for a new approach to COVID diagnosis.

The strong correlation of Ag concentrations with Ct values, and the high sensitivity of the S-PLEX Ag test in samples covering the range of viral loads observed in newly symptomatic individuals with COVID-19, suggests that the high false negative rates that have been observed with Ag RDT tests (versus PCR) in similar populations are due to the relative analytical insensitivity of the RDT tests, rather than any major difference in the kinetics of expression of Ag and RNA in the nose/nasopharynx. A highly sensitive assay could detect individuals earlier in infection than an Ag RDT or, potentially, support Ag testing in pooled samples.

Studies such as ours that use a high sensitivity quantitative assay to measure an infectious disease biomarker provide a useful dataset for predicting the clinical performance of less sensitive methods, or for setting analytical sensitivity targets to achieve specific clinical performance targets. To this point, our estimated clinical sensitivities for the BD and Quidel RDTs, based on our measured distributions of Ag concentrations, are consistent with the field performance of Ag RDTs in a recent European prospective study [3]. The report evaluated Ag RDT test performance at POC versus PCR and found that the best-performing visually-read Ag RDT was 76.6% sensitive and 99.3% specific, with high sensitivity in samples with Ct values < 25, moderate sensitivity in samples with Ct <30, and poor sensitivity above Ct 30 [3].

One important consideration for COVID-19 diagnostics is their ability to identify patients with high loads of active virus that are more at risk of transmitting disease. Several investigators have reported difficulty in culturing virus from patient samples with measured viral loads below approximately 1×10^5^ RNA copies/mL [21-24]. However, virus has been recovered from samples with RNA levels as low as 1.2×10^4^ copies/mL [25], and from samples with a Ct value of 34-35 on a range of PCR assays [26-28]. Using our conversion factor from the correlation of Ag and RNA levels, 1 x 10^4^ copies/mL translates to 1.5 pg/mL, which is about 5 times higher than the S-PLEX Ag assay cutoff (0.32 pg/mL), suggesting that the S-PLEX Ag assay should be able to detect Ag in most if not all samples from which virus is culturable (though we note cultures themselves have variable sensitivity [29], and that lack of ability to culture virus doesn’t preclude transmission).

If converted to a clinical diagnostic, the S-PLEX Ag assay offers some potential advantages over existing diagnostic approaches. The assay’s analytical sensitivity is similar to that of PCR-based methods, and the assay was able to detect nearly all samples with a Ct of ≤ 35. The S-PLEX Ag assay is approximately 500-fold more sensitive than existing Ag RDTs with electronic readers, with near perfect specificity in clinical samples in UTM and normal saline. We found a higher rate of false negatives for samples having Ct values between 36 and 38 than would be expected based on the correlation of Ct values with Ag concentration. These false negative samples were largely pediatric samples eluted in saline. It is possible that there may be some loss of Ag by adsorption from saline, when compared to samples eluted in protein-containing UTM, warranting further study. Depending on the use case, the inconsistent detection of samples with Ct values > 35 could be a disadvantage (if the goal is to detect all individuals with evidence of infection, e.g. to optimize hospital safety and inform contact tracing), or an advantage (if the goal is to identify those most likely to transmit [29]). The high specificity of the S-PLEX Ag assay, if confirmed in a clinical diagnostic setting, should be an advantage over Ag RDTs, some of which have generated concerning specificity data in actual practice [8] despite having high specificities in EUA validation studies [5, 6].

While the S-PLEX Ag test offers analytical advantages over Ag RDTs, given that the Ag RDTs can be performed at POC, the operational characteristics of the S-PLEX Ag assay (cost, speed, operator time) must be carefully considered [30]. The turnaround time of the assay is 4 hours, and 78 samples can be run on each plate. A single operator can process as many as 5 plates (390 samples) in a run, with higher throughputs available with automation. The assay utilizes entirely different reagents than used for molecular testing, which might make it less susceptible to current supply chain issues.

Our study has some limitations. First, this was a retrospective study using frozen samples previously tested by PCR; future studies will test fresh samples prospectively, but the tight correlation between Ag and Ct values suggests that freezing was not a significant issue. We note that the sensitivity in the pediatric sample set was lower than in the adult sample set, but ascribe this to a higher Ct value distribution in the pediatric samples and, possibly, to a higher proportion extracted in saline relative to UTM. For some of the patients who provided samples, the time between onset of symptoms and testing was long, but this reflects real life clinical test use. The majority of the patients whose samples were included in our study were symptomatic, so the results will need confirmation in asymptomatic individuals [31].

In summary, we have shown that an ultrasensitive Ag assay is able to detect Ag throughout the range of viral load in clinical samples in adults and children infected with SARS-CoV-2. The assay has high sensitivity and specificity, offering an alternative to PCR and a clear analytical advantage over Ag RDT. Future prospective studies evaluating test performance in programmatic screening of symptomatic and asymptomatic individuals will demonstrate whether the S-PLEX Ag assay can offer a diagnostic alternative that is inexpensive, rapid, and high-throughput, thus contributing a novel diagnostic tool for management of this pandemic.

## Data Availability

All data are available upon request

## Acknowledgments

We thank Richard Rossi for assistance with PCR Ct data capture and all of the virology laboratory staff from Boston Children’s Hospital for their assistance with sample aliquoting. We thank Pradeepthi Bathala, Shraddha S. Kale, Salvia Misaghian, Nikhil Padmanabhan, Daniel Romero and Navaratnam Manjula for conducting the assay measurements. The following reagent was deposited by the Centers for Disease Control and Prevention and obtained through BEI Resources, NIAID, NIH: SARS-Related Coronavirus 2, Isolate USA-WA1/2020, Gamma-Irradiated, NR-52287.

