## Supplemental Material for "Correlation of SARS-CoV-2 nucleocapsid antigen and RNA concentrations in nasopharyngeal samples from children and adults using an ultrasensitive and quantitative antigen assay"

**Supplementary Materials**

**Methods**

Immunoassay Protocols

Both U-PLEX® and S-PLEX® formats follow similar processes for the initial antibody binding steps: (i) incubating a biotin-labeled capture antibody in the wells of an MSD® U-PLEX 96-well plate (R-PLEX assay), or MSD GOLD™ streptavidin 96-well plate (S-PLEX assay) to immobilize the capture antibody; (ii) adding the sample and an equal volume of the kit Assay Diluent to the wells and incubating to bind analyte to the capture antibodies; (iii) adding detection antibody labeled with the SULFO-TAG™ ECL label (R-PLEX assay) or TURBO-BOOST™ label (S-PLEX assay) and incubating to bind captured analyte. The S-PLEX format involved two additional signal steps that use the TURBO-BOOST label to generate an enhanced signal: (iv) incubation with MSD Enhance buffer and (v) incubation with TURBO-TAG™ Detection Solution. Each incubation step was carried out on a plate shaker (700 rpm) at room temperature, except for the TURBO-TAG incubation, which was carried out in a heated shaker at 27$^{o}$C. Each incubation step was followed by a washing step on an automated 96-well plate washer to remove excess reagent or sample.

To detect the bound SULFO-TAG label in the R-PLEX format or TURBO-TAG detection reagent in the S-PLEX format, an ECL read buffer (MSD GOLD Read Buffer B) was added to each well and the plates were analyzed on an MSD® ECL plate reader (MESO SECTOR® S 600, SECTOR Imager 6000, or MESO QuickPlex® SQ 120). The readers apply electrical potentials to the electrodes in the plates and use a sensitive CCD camera to image and quantify the induced ECL signals.

Both assay formats require 25 μL of sample per measurement. Upper respiratory swabs eluted into viral transport media or PBS can be analyzed without any additional sample preparation. Swab elution media containing protein denaturants – including proprietary media provided with certain molecular assay kits such as the Hologic Specimen Transport (HST) media – may interfere with the assay and should be avoided. The R-PLEX format has a total assay time of about 4 hours (or 3 hours if batches of plates with immobilized capture antibody are prepared in advance). The S-PLEX format has a total assay time of about 5 hours (or 4 hours if the capture antibody is immobilized in advance). For both, hands-on time is approximately 1 hour.

Immunoassay Quantitation and Analysis

To calculate the concentration of nucleocapsid in the samples, an eight-point calibration curve with recombinant SARS-CoV-2 nucleocapsid protein (full length with c-terminal His6 epitope tag expressed in mammalian cells; provided with the assay kits) in the kit Assay Diluent was run in duplicate on each plate. The calibration assay signals as a function of protein concentration were fit to a four parameter logistic (4PL) curve using $1/Y^2$ weighting. Nucleocapsid concentrations in test samples were determined by back-fitting the measured assay signals to the 4PL curve. In addition to the calibration samples, high and low concentration control samples were run in duplicate on each plate to ensure consistent quantitation.

Limits of detection (LODs) were determined as the concentration of recombinant nucleocapsid that provides a signal at least three standard deviations above the assay background generated using the kit Assay Diluent as the sample (as determined by back-fitting to the 4PL calibration curve). For assessing sample positivity, we set a relatively conservative threshold of 2 x LOD.

Spike Recovery Experiments

Spike recovery experiments were conducted on some of the clinical samples to evaluate the potential for matrix interference. In these experiments, two measurements were made per sample: (i) a baseline measurement conducted using the standard assay protocol and (ii) a spiked measurement conducted by spiking 50 pg/mL recombinant nucleocapsid into the assay diluent during the sample incubation step. Percent recovery was calculated as the measured concentration in the spiked sample, minus the measured concentration in the baseline sample, divided by 50 pg/mL.

<u>Statistical analysis</u>

Graphs were generated using TIBCO Spotfire Desktop 10.6.1. Statistical analyses were carried out using the R statistical computing language (version 3.6.2). Confidence limits on proportions were calculated using the binom.test function. Linear regression was calculated using the lm function. Receiver operating characteristic (ROC) curve area under curve (AUC) values and confidence intervals were calculated using the pROC library in R. Sensitivity and specificity were calculated using clinical PCR results as the reference method. Confidence intervals were calculated to 95% confidence (95%CI).

**Supplementary Table 1**
**Samples with positive PCR but negative Ag test results**

| Adult vs Pediatric | Days from onset of symptoms | Nucleocapsid concentration (pg/mL) | PCR Ct value |
|---|---|---|---|
| Adult | 23 | < 0.2 | 37.4 |
| Adult | 5 | < 0.2 | 39.6 |
| Adult | 0 | < 0.2 | 38.0 |
| Ped | 14 | < 0.2 | 34.5 |
| Ped | 14 | < 0.2 | 38.4 |
| Ped | 4 | 0.3 | 36.7 |
| Ped | 4 | 0.2 | 38.1 |
| Ped | 28 | < 0.2 | 37.2 |
| Ped | 2 | 0.3 | 30.3 |
| Ped | 3 | < 0.2 | 36.4 |
| Ped | 39 | < 0.2 | 33.3 |
| Ped | 5 | < 0.2 | 38.2 |
| Ped | 3 | < 0.2 | 38.0 |
| Ped | 2 | < 0.2 | 37.7 |
| Ped | 1 | 0.2 | 36.6 |
| Ped | 5 | 0.3 | 36.8 |
| Ped | 3 | < 0.2 | 37.4 |
| Ped | 3 | 0.3 | 38.2 |
| Ped | 2 | < 0.2 | 38.3 |
| Ped | 1 | < 0.2 | 37.2 |
| Ped | 3 | < 0.2 | 37.1 |
| Ped | 7 | < 0.2 | 37.9 |
| Ped | 3 | < 0.2 | 37.7 |
| Ped | 7 | 0.2 | 36.3 |

**Supplementary Table 2**
**Performance of the R-PLEX Ag assay versus PCR for detection of SARS-CoV-2 in samples from adult and pediatric patients**

| | PCR-Negative Patients | | PCR-Positive Patients All Ct Values | | Ct Values ≤ 35 | |
|---|---|---|---|---|---|---|
| | N | Specificity (95% CI) | N | Sensitivity (95%CI) | N | Sensitivity (95%CI) |
| **Age Group** | | | | | | |
| Pediatric | 40 | 100% (91%-100%) | 101 | 67% (57%-76%) | 72 | 89% (79%-95%) |
| Adult | 50 | 100% (93%-100%) | 35 | 77% (60%-90%) | 29 | 93% (77%-99%) |
| Combined | 90 | 100% (96%-100%) | 136 | 70% (61%-77%) | 101 | 90% (83%-95%) |

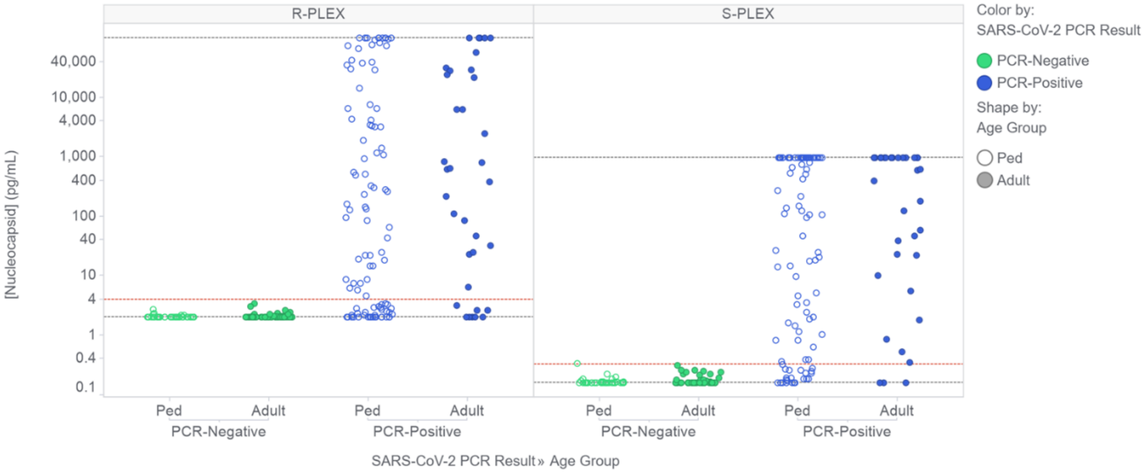


**Supplementary Fig 1**. Measured nucleocapsid concentrations in clinical NP swab samples from PCR-negative (green) and PCR-positive (blue) adults (filled circles) and children (Ped, open circles). The figure shows the concentrations measured with the conventional (R-PLEX, left panel) and ultrasensitive (S-PLEX, right panel) ECL assays on undiluted samples. Concentrations above the highest calibration standard or below the limit of detection (dashed grey lines) were assigned those values, respectively. The dashed red line indicates the assay thresholds for classifying samples as positive.
